# Understanding Post-Surgical Recovery in Vestibular Schwannoma: A Qualitative Exploration of Patient Experiences

**DOI:** 10.64898/2026.01.09.26343681

**Authors:** Nora Nieke, Lisa Brandes, Svenja Wandke, Isabelle Scholl, Mareike Rutenkröger

## Abstract

**Introduction:** Vestibular schwannoma (VS) is a benign tumor of the vestibulocochlear nerve, often causing hearing loss, balance disturbances, and psychosocial challenges. While surgical resection is standard, the long-term biopsychosocial impact of surgery is poorly understood.

**Research question:** What are the physical, psychological, and social challenges experienced by patients up to five years after VS surgery?

**Material and Methods:** A qualitative study was conducted using semi-structured video call interviews with 12 patients recruited via a patient advocacy group. Interviews explored postoperative experiences across physical, psychological, and social domains. Transcripts were analyzed using thematic content analysis with a coding system developed deductively and refined inductively. Data saturation was reached after 12 interviews.

**Results:** Participants reported diverse physical symptoms, including hearing loss, tinnitus, dizziness, pain, fatigue, and facial nerve palsy. Psychological challenges included anxiety, depression, cognitive difficulties, and reduced stress tolerance. Social changes encompassed strained relationships, withdrawal from work and leisure activities, and limited social participation. Physical, psychological, and social challenges interacted dynamically, with emotional distress amplifying social isolation and healthcare provider support influencing coping and adaptation.

**Discussion and Conclusion:** VS surgery has a multifaceted, long-term impact on patients’ lives. The interplay of physical, psycho-logical, and social challenges underscores the need for holistic, multidisciplinary care, early patient education, and integration of supportive interventions. Engagement from healthcare providers plays a key role in mitigating distress and facilitating adaptation. These findings highlight the importance of addressing biopsychosocial aspects to improve long-term recovery and health-related quality of life in VS patients.

## 1. Introduction

Vestibular schwannoma (VS), also known as acoustic neuroma, is a benign tumor that typically arises on the vestibulocochlear nerve, impacting both hearing and balance [1]. VS represent approximately 8% of all intracranial tumours and, at the same time, they are the most common intracranial tumours in the cerebellopontine angle in adults, accounting for 85% of cases [1]. While surgical removal of the tumor is the primary treatment option, the postoperative phase often presents significant challenges [2–5]. Patients frequently experience a range of physical symptoms, such as hearing loss, balance disturbances, and fatigue, alongside psychological difficulties including anxiety and depression [6–10] and social challenges [11]. Despite the medical focus on surgical outcomes and recovery, there is limited qualitative research exploring the broader biopsychosocial impact of VS and its treatment on patients’ lives.

While previous studies have addressed individual aspects of recovery, a comprehensive understanding of the combined biological, psychological, and social difficulties faced by patients remains under-explored. This gap in knowledge is critical, as a holistic approach to post-surgical care can enhance rehabilitation and improve overall health-related quality of life (hrQoL) for patients.

## 2. Research Question

The aim of this study was to explore the biopsychosocial challenges faced by individuals following VS surgery. Specifically, we sought to answer the research question: What are the biological, psychological, and social challenges experienced by patients up to 5 years after VS surgery?

## 3. Methods

### 3.1 Study design

A qualitative study was conducted using semi-structured video call interviews, guided by the Consolidated Criteria for Reporting Qualitative Research (COREQ) [12], see Suppl. I.

### 3.2 Recruitment

Recruitment was facilitated through the patient advocacy group Vereinigung Akustikus Neurinom e.V. (VAN). Recruitment through a patient advocacy group may introduce selection bias, as individuals in such groups may be more likely to experience pronounced postoperative challenges. NN presented the study objectives at a national group meeting, and a flyer detailing the study was distributed across VAN’s regional groups starting on January 26, 2024. Initially, 18 potential participants had expressed interest. Eligibility criteria included surgical removal of the tumor, surgery performed within the last 5 years, and experiencing biopsychosocial challenges post-surgery. These criteria were verified during an initial telephone screening.

### 3.3 Procedures

The study received approval from the local Ethics Institutional Review Board (LPEK-0694). Participants were invited to take part via email, with the requirement that they have access to a device with an internet connection for participation. Written informed consent was obtained from all interested participants prior to enrollment. Demographic data were collected through questionnaires. Descriptive data were analyzed using SPSS software. Subsequently, participants were contacted via email to schedule the semi-structured interviews, which were conducted by NN between February 19, 2024, and March 30, 2024. All videocall interviews were recorded using a digital tape recorder, with the average interview lasting 54 minutes. The recordings were transcribed verbatim by NN for further analysis.

### 3.4 Interview Domains

The semi-structured interview guide was developed using open-ended questions to elicit information about the biopsychosocial challenges faced by participants during the postoperative period (<5 years after surgery). The preliminary interview guide was reviewed and refined with input from two VAN advocates, resulting in in a modified final version with improved clarity, comprehensibility and relevance. The primary goal was to explore a broad range of topics related to the (1) physical, (2) psychological, and (3) social changes experienced by participants following surgical treatment, ensuring a comprehensive understanding of their postoperative experiences.

### 3.5 Data Analysis

The data analysis involved coding the full text of twelve transcribed interviews using MAXQDA (Version 2024). Following Kuckartz [13], meaningful sections were used as units of coding, with a meaningful phrase as the minimum and a multi-phrase text segment as the maximum unit. Multiple codings were applied when necessary. The coding system is divided into two hierarchical levels and includes three main categories, and 16 subcategories (see Suppl. II). Based on the content structure of the interview guide, the coding system was first divided into three deductive main categories: 1) physical changes, 2) psychological changes, and 3) social changes. NN (first author) then carried out an inductive categorization based on the entire data material. The focus was on identifying and analyzing how physical, psychological, and social changes interacted with each other, with particular attention to the causal relationships and reciprocal influences described by participants. The preliminary version of the coding system was revised with MR and SW. NN subsequently coded the entire data set a second time using the final coding system, LB recoded the data set. Conflicts were settled through consensus discussion with MR. Data saturation was reached when no new relevant themes emerged from the interviews. Transcripts were not returned to participants due to the exploratory nature of this study.

## 4. Results

### 4.1 Demographic and Clinical Characteristics

The sample consisted of N=12 participants, ranging in age from 27 to 70 years (M=49.58; SD=11.88). Seven participants identified as female. Most had more than 11 years of schooling (75%) and were employed (83.3%). Time since diagnosis varied, with the largest group diagnosed four years earlier (41.7%). Half of the participants had received treatment 2–3 years ago. Tumour size was most commonly classified as Koos grade 2 (50%). Retrosigmoidal surgery was the predominant treatment approach (83.3%). Seven participants (58.3%) used a hearing aid. Preoperatively, the most frequent symptoms were acute hearing loss and hearing impairment (each 50%), followed by dizziness (41.7%). Postoperatively, the most common symptoms were hearing loss (83.3%), tinnitus (75%), dizziness (75%), and headache (66.7%), along with notable rates of balance disorder (41.7%) and fatigue (66.7%). Full sample characteristics are reported in Table 1.

**Table 1.**
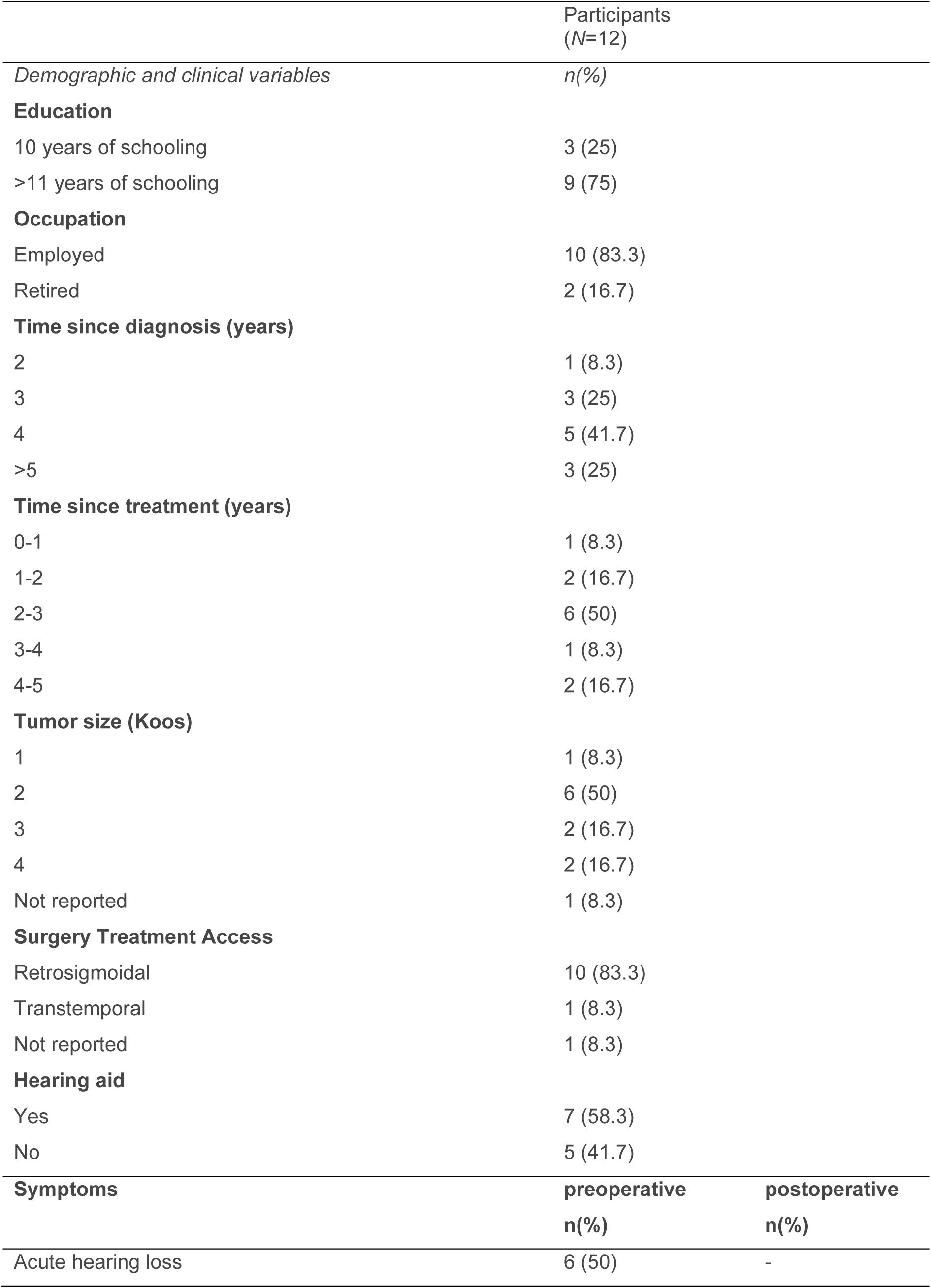
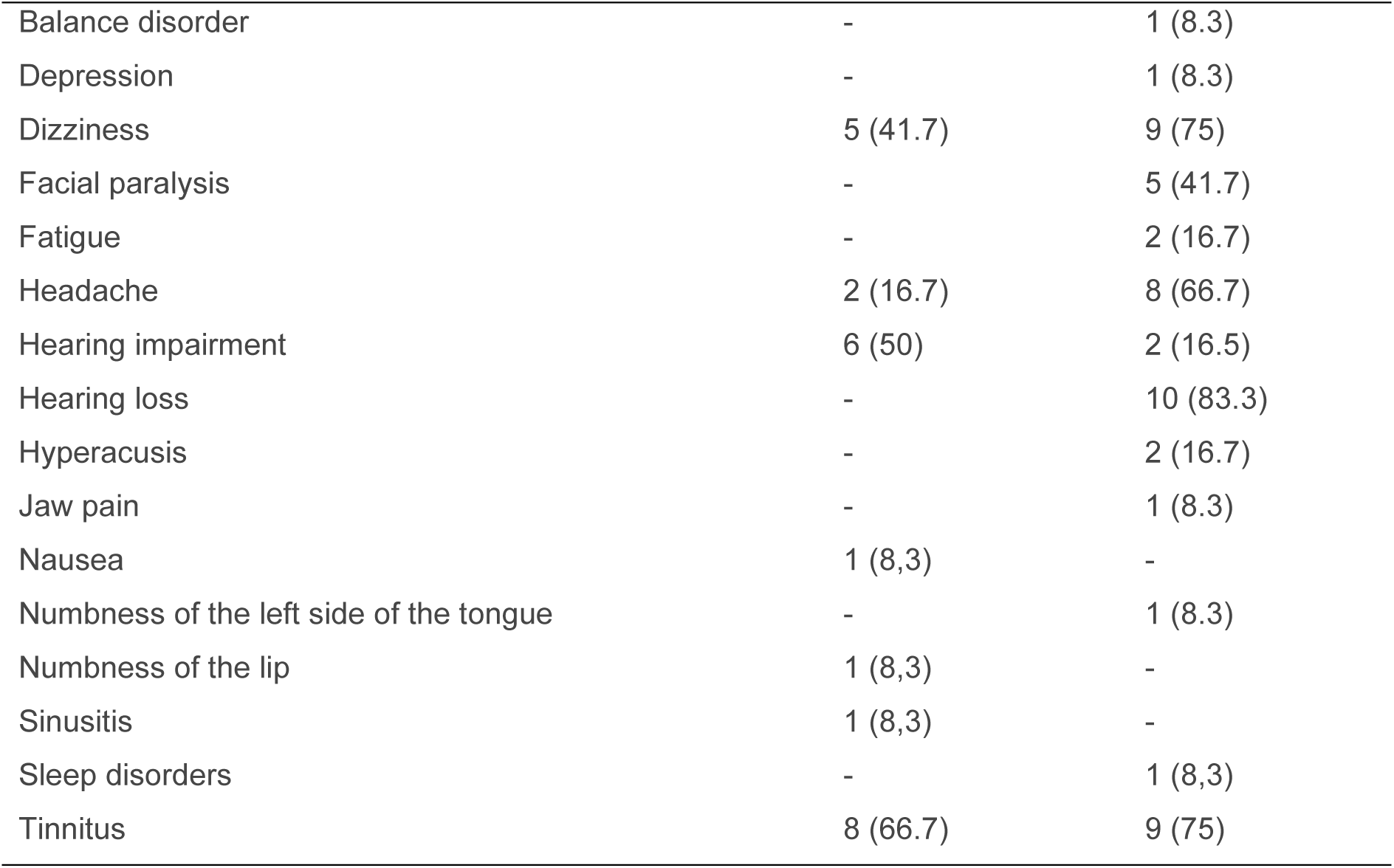
Sample characteristics.

### 4.2 Themes

The following paragraphs elaborate on the deductively defined themes. Tables 2, 3, and 4 provide a structured overview of the four themes, including relevant subthemes (where applicable) and corresponding participant quotes.

**Table 2.** Theme 1: Physical Changesi.

| Subthemes | Data extracts |
| --- | --- |
| Altered hearing | "Because I simply want to protect myself physically and I also know that the tinnitus number is then very quickly not the physical number, but rather a burden on the psyche, because you are constantly this noise with the pink elephant in the room and that's just very annoying."(ID1)<br>"And (.) I experienced the deafness as totally dramatic at the beginning. I was also told in the information session that it can happen, but most people are fine with it. I couldn't cope with it at all. It drove me completely crazy. Always hearing the noises from the wrong side." (ID13) |
| Dizziness | "Immediately afterwards, you're a complete mess, so that's the short version: headache, vomiting and dizziness for the first two or three days. I think I got up again after three days with help." (ID2) |
| Pain | "I actually had a lot of problems with my jawbone. On the opposite side, i.e. the left side. This pressure with the scar (.) only gradually smoothed it out a bit with manual therapy. I really had pain in my jaw joint here." (ID1)<br>"Where I have struggled for a long time - or where I still struggle a bit today - is that there is of course a fairly thick cut across the head into the neck. And that often leads to tension for me. It causes pain at the scar." (ID10) |
| Fatigue | "And then there's the permanent exhaustion. So a permanent massive exhaustion." (ID16) |
| Facial paralysis | "After I was allowed back into the bathroom for the first time, I thought: "The eye is staring straight ahead as if it's no longer alive.". That was creepy. And the left (.) half of the face was completely drooping. The eye was hanging down. The mouth was hanging down." (ID8) |
| Other symptoms | "I have a numb left tongue. It is still numb. Partly also flavor- how should I put it? Tasteless, so I can't taste it very well. It always goes over like this, left right half of the tongue. When I ate bread straight after the operation and no matter what I ate, I didn't like the taste." (ID11)<br>"So I couldn't sleep. No medication worked. They gave me something to sleep on and I didn't feel like I was sleeping at all. I was so tense I was there." (ID9) |

**Table 3.**
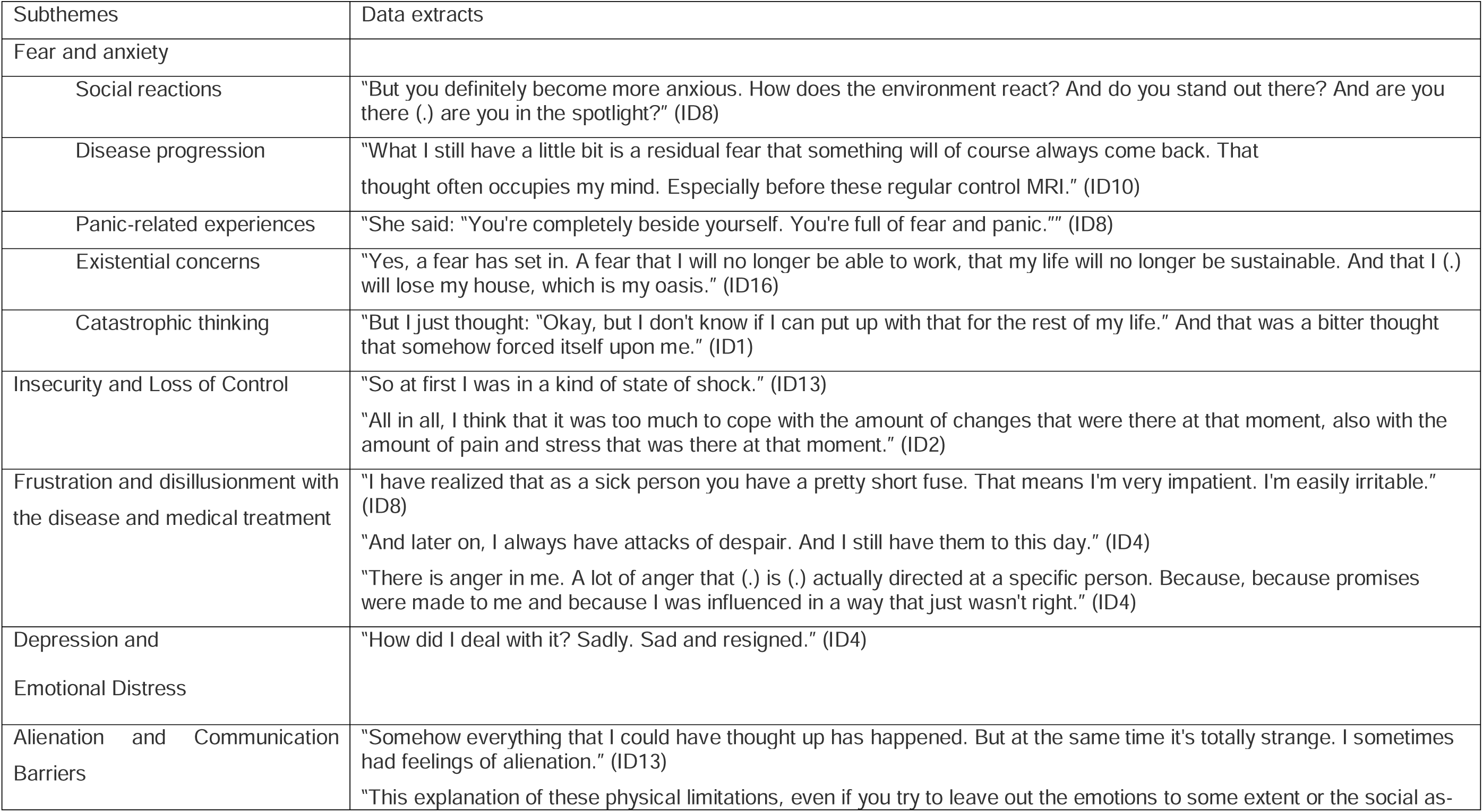
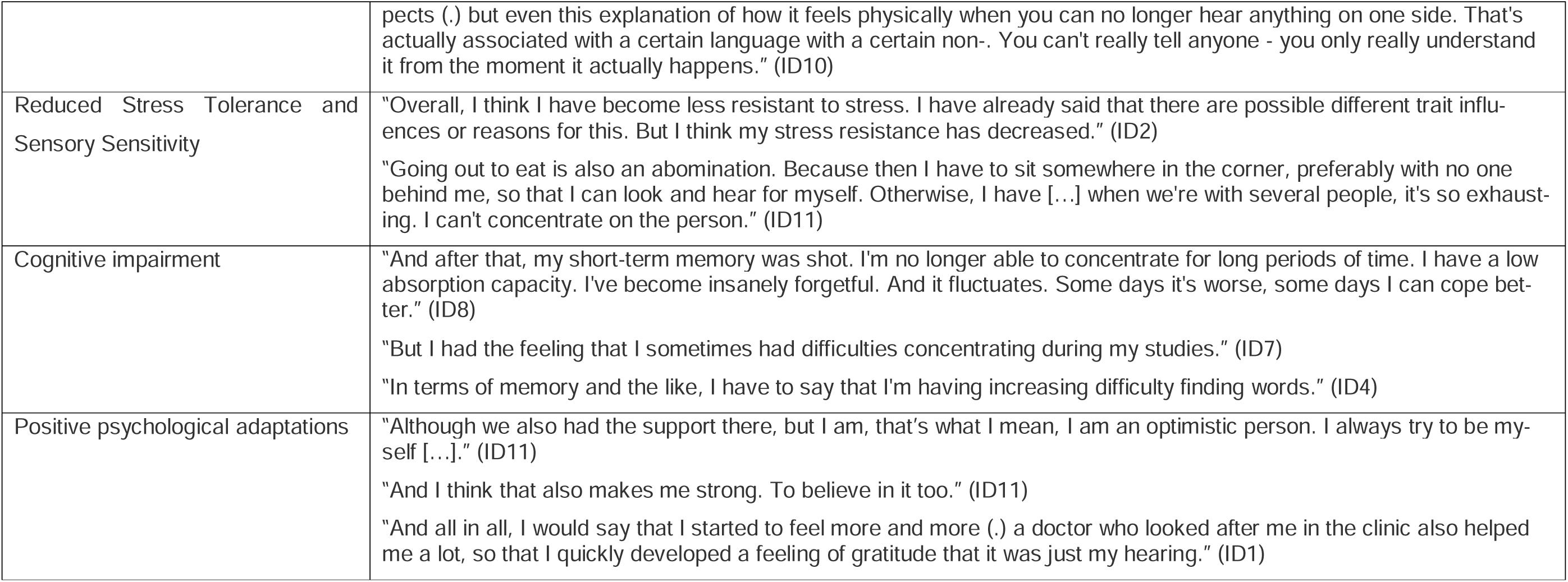
Theme 2: Psychological Changes.

**Table 4.** Theme 3: Social Changes.

| Subthemes | Data extracts |
| --- | --- |
| Social interactions |  |
| Strain on the family and/or partnership | "I have these moments when I like to be alone sometimes and, as I said, I don't want to talk to anyone or something (.). That has perhaps become a bit more. And it may well be that (1) family, friends and partners suffer a bit as a result." (ID12) |
| Withdrawal from social contacts | "Many friends have withdrawn. Many doors have closed." (ID8) |
| Confusion and/or shock | "I also had the feeling that friends were calling to check "How is she mentally now? Is she completely confused? Or does she (I'm exaggerating here) still have all her wits about her? Or is she anything like she used to be?"." (ID8)<br>"After you might take a short walk around the factory premises after lunch and then I make a very strange movement, passing behind the person to get from the right side to the left side. It irritates most people at first and then you have to explain." (ID2) |
| Acknowledgment of open communication | "I think I always thought it was important that the people around me knew what was going on. And of course they react completely differently to it, but in my environment it's almost always positive." (ID10) |
| Changes in social participation |  |
| General interaction difficulties due to altered hearing | "Of course I have that now too. When I'm in an environment where it's very loud, I sometimes find it difficult (.) to understand the person I'm talking to." (ID10) |
| Workplace accommodations | "Professionally, I started at the same time that I had been given a severely disabled status of 30% or 30 points. So it was clear that my employer would actually put me on an equal footing." (ID10)<br>"It was important to me that (.) I now have a particular place where I always sit because it's the quietest place we have and my ear is to the window and not where someone on the left could approach me from the corridor. I've decided for myself that I'm going to demand things the same way and definitely not differently. And there's no need to argue about it. Of course, it was much easier when I was on an equal footing." (ID1) |
| Restrictions on leisure activities | "One social change was that I could no longer take part in so many things.<br>I could take part in." (ID10) |

#### 4.2.1 Theme 1: Physical Changes

##### 4.2.1.1 Altered hearing

All participants reported altered hearing symptoms, including tinnitus, unilateral hearing loss, hyperacusis, and binaural hearing impairment. Tinnitus was described as particularly noticeable during stress or exhaustion, and some participants reported it as mentally exhausting. Four participants reported permanent tinnitus, while two ITs did not perceive any tinnitus at times. ID1 established a link between the physical stress caused by the tinnitus and her psychological well-being: “*Because I simply want to protect myself physically and I also know that the tinnitus number (.) is then very quickly not the physical number, but rather a burden on the psyche, because you constantly hear this noise with the pink elephant in the room and that is simply very (.). The worst thing about this tinnitus is really that I (.) am so cognitively challenged that I can’t concentrate on much at the same time. And that’s just extremely exhausting and I just don’t feel like it. And then I get into such a rut again that I feel very sad and depressed.”*

Unilateral hearing loss, caused by surgical nerve transection, led to disorientation and stress. While two ITs initially did not notice the hearing loss at all, ID13 reported considerable difficulties in dealing with the unilateral deafness: “*And (.) I experienced the deafness as totally dramatic at the beginning. I was also told in the information session that it can happen, but most people cope well with it. I couldn’t cope with it at all. It drove me completely crazy. Always hearing the noises from the wrong side. […] And it drove me totally crazy when the cars always came from the wrong side, so to speak. When I looked in the wrong direction. I found it extremely stressful (to the point of being traumatized?), the one-sided deafness.”*

Another physical change mentioned was hyperacusis, which manifests itself in the form of severe overstimulation in the cognitive processing of acoustic stimuli. The ITs said that they found everyday situations, such as traffic noise, listening to music or following a conversation in a large group, challenging. One participant with pronounced hyperacusis described her experience as follows: “*For example, if I were to go into [electronics shop] now to buy some device. The frequencies that come from this (.) from this building, from these many televisions that are on. I (.) collapse on the spot. They are electric shocks in my head.”*

##### 4.2.1.2 Dizziness

Participants experienced dizziness, including balance issues, nausea, and mild to rotational dizziness. Balance problems often persisted after surgery, especially in low-light conditions or with rapid head movements. Three different types of dizziness were mentioned, with the most common being temporary mild dizziness following surgery, often compared to a minimally intoxicated state. One participant reported severe spinning vertigo after surgery. ID2 explained his symptoms after the operation as follows:

*“Immediately afterwards you’re completely fucked up, so that’s the short version: headache, vomiting and dizziness for the first two or three days. I think I got up again after three days with help.”*

##### 4.2.1.3. Pain

Pain was categorized into postoperative headaches, scar pain, and jaw pain. Some participants experienced acute headaches post-surgery, while others had persistent pain. Scar pain sometimes triggered migraines, and jaw pain appeared months or years later.

##### 4.2.1.4 Fatigue

Some participants described pronounced fatigue, which either occurred only in the first few days or weeks after the operation or remained a constant burden. One participant noted that, since returning to work and being exposed to increased stress, she could walk for a maximum of 15-20 minutes before becoming exhausted.

##### 4.2.1.5 Facial nerve palsy

Unilateral facial nerve palsy was reported by five participants. These participants were particularly distressed by the altered appearance of their face, which led to irritated looks and comments from their social environment. One participant described her first impression following the operation as follows: *After I was allowed back into the bathroom for the first time, I thought, “That eye is staring straight ahead as if it’s not alive at all. That was scary. And the (.) left side of my face was completely drooping. The eye was hanging down. The mouth was hanging down*.

##### 4.2.1.6 Other symptoms

A few participants reported sleep disturbances. One participant described synkinesia on the affected side of the face due to excessive facial training. He explained that the corner of his mouth was pulled up when he closed his eyes, accompanied by painful cramps in the affected side of his face at night. Another participant developed hyperkinesia, which she attributed to improper facial nerve palsy treatment using the Proprioceptive Neuromuscular Facilitation method. One participant reported scalp irritation as a rare physical change, while another experienced tongue numbness immediately after surgery, accompanied by a loss of taste, although it subsided over time.

#### 4.2.2 Theme 2: Psychological Changes

Participants reported a range of psychological challenges. Their experiences encompassed fear, insecurity, frustration, depressive symptoms, alienation, reduced stress tolerance, cognitive impairments, psychiatric disorders, and, in some cases, positive psychological adaptations.

##### 4.2.2.1 Fear and Anxiety

Fears were categorized into five domains: social reactions, disease progression, panic-related experiences, existential concerns, and catastrophic thinking. Some participants feared social stigma, being perceived as “different,” or receiving unwanted special treatment. Distress was often triggered by medical procedures, such as MRI scans, or physical limitations, particularly in relation to vertigo and concerns about losing independence. ID10 described the fear of progression with the words: “*[…] what I still have a bit of is a (..) residual fear that something will of course always come back. I often think about that. Especially before these regular check-up MRIs. I would say that’s a psychological burden. Just the fear that the whole (.) that there’s something there again and the whole thing will start all over again.”*

Anxiety was exacerbated by postoperative complications, financial insecurity, and potential job loss. Several participants engaged in catastrophic thinking, fearing worsening health, repeated surgical interventions, or complete hearing loss, leading to significant emotional distress. ID8 reported experiences in hospital that had caused her increased anxiety and panic. She had to undergo emergency surgery after the operation due to a backlog of cerebrospinal fluid and was subsequently discharged from hospital with a swelling on her abdomen. After the practitioners told her that something like this could happen at any time, she anxiously began to examine her body regularly. Her occupational therapist commented: “*You are completely beside yourself. They are full of fear and panic.*” (ID8).

ID1 had a particularly impressive experience of the catastrophizing thought of also going deaf in the opposite ear: *“And I remember exactly that I (..) the worst psychological event was actually […] six weeks after the operation, I would say. I was looking at an exhibition in a castle and suddenly thought: […] “What if I lose the other hearing on my left side as well?”. And that was so exuberant. […] And there was something very frightening about that. Because I’m already a very positive person and it had something of (…) I didn’t want to kill myself at the moment when I thought that. But I just thought: “Okay, but I don’t know if I can bear that for the rest of my life.” And that was a bitter thought that somehow forced itself on me.”*

##### 4.2.2.2 Insecurity and Loss of Control

Half of the participants described profound feelings of insecurity, including initial shock in the weeks after surgery, and a loss of autonomy. Some felt overwhelmed by environmental stimuli, particularly noise, or powerless when attempting to explain their hearing loss to others. Chronic pain contributed to a sense of hopelessness, leading to social withdrawal and the abandonment of previously enjoyed activities. The realization of the severity of their diagnosis further reinforced feelings of despair, e.g. due to the constant pain, ID4 had become “*much quieter*”, no longer took part in leisure activities and felt excluded from her social environment. She described her situation as follows: *“I used to love flea markets. All these things have been denied to me. And (.) I am, we built ourselves a mobile home. Now that I’m retired, I wanted to see the whole world. And I can hardly get into town because I can’t stand the rocking in the car any more. All these things make me desperate.”*

##### 4.2.2.3 Frustration and disillusionment with the disease and medical treatment

Participants expressed frustration related to persistent headaches, prolonged waiting times in medical settings, and reduced tolerance for stress. Some were dissatisfied with perceived false reassurances from medical professionals and the lack of psychological support, leading to anger and disappointment. Others experienced a sense of disillusionment, believing that their illness was an isolating burden that could not be fully understood or shared by others. One participant associated her lower frustration tolerance with the disease: “*I’ve noticed that as a sufferer you have a pretty short fuse. That means I’m very impatient. I’m easily irritable. I don’t know all that. I used to be such a sweet, patient little sheep that you could do anything with.”*

ID4’s frustration was combined with her accusations towards the surgeon. Her anger was directed in particular at the surgeon, who had made false promises to her and said: “*Mrs. XY, after six weeks you can forget about the disease*.” She emphasized that he had also reassured her with the words “*Don’t worry Mrs. XY. If anyone can do it, it’s you!*” (ID4) to reassure her. In addition, he had put pressure on her and said that she had to make a decision as quickly as possible. She also expressed her dissatisfaction to other practitioners: “*But basically they all talk, yes, how can I say that? I should do autogenic training, I should (.) I should do progressive muscle relaxation. None of that helps me with the headache. It simply doesn’t help. And then, when I’m told that again and again, I say to myself: “I want you to have my headache for an hour. My head for an hour. And then say that again.”*

##### 4.2.2.4 Depressive symptoms and Emotional Distress

Half of the participants reported experiencing depressive symptoms, with contributing factors including workplace bullying, social isolation, and difficulty coping with their diagnosis. Feelings of sadness were particularly pronounced in those who had lost their familial roles or lacked adequate support following surgery. Several participants described increased rumination, engaging in deep reflection about their condition and life circumstances, particularly when experiencing persistent symptoms such as chronic headaches.

##### 4.2.2.5 Alienation and Communication Barriers

Three participants reported profound feelings of alienation, describing sensations of detachment or unreality. ID13 also felt as if he had been torn out of his old reality and reported: “*And what came as a particular psychological symptom was that I sometimes had the feeling that I no longer understood the whole world, the whole world seemed to me totally (.) somehow no longer, no longer like the world I know. I always had the feeling that I was in such a (.) in a bad dream. I’m under anesthesia, I’m waking up now. […] None of this can be true. […] I had such feelings of alienation sometimes.”*

Some struggled to communicate their experiences effectively, feeling that those with full hearing could not comprehend the challenges of unilateral deafness or other invisible consequences of their illness. For example, ID1 (47) said that she reached her limits when explaining her physical limitations, i.e. when she tried to explain to others *“how it feels physically when you can no longer hear on one side”.* She described how she no longer knew what it felt like to hear on both sides from the time she became deaf on one side: “*It’s actually with a certain speech- (.) with a certain non- (.). You can’t really tell anyone - (.) you only really understand it from the moment it actually happens. And that’s why you know that no one will really be able to understand it. That’s also simply a difficulty. And you know it’s also very strange, but it’s also good. You immediately no longer know what stereo hearing feels like. That’s why you know that someone who can hear stereo won’t be able to understand what it’s like (.) what kind of physical limitation you have.”*

##### 4.2.2.6 Reduced Stress Tolerance and Sensory Sensitivity

Participants commonly reported decreased resilience and increased fatigue, with some describing a persistent state of “emergency mode” as they attempted to manage daily life. Loud and chaotic environments were particularly overwhelming, with difficulties intensifying in situations where multiple people were speaking simultaneously. Sensitivity to noise was a recurrent issue, with one participant (a teacher) noting that classroom noise exacerbated headaches and stress. Noisy environments with a large number of acoustic stimuli were a particular challenge for participants. ID1 described it as follows:

> “I think I was very sensitive to noise anyway, I was before, but it had another dimension to it. At first it was very difficult for me to talk to anyone and still enjoy it when it was loud around me.”

##### 4.2.2.7 Cognitive Impairments

Cognitive difficulties were prevalent, with concentration problems reported by six participants. Many described feeling mentally exhausted during conversations or struggling to maintain focus on tasks. Participants also reported memory deficits, including short-term memory loss and word-finding difficulties.

##### 4.2.2.8 Positive Psychological Adaptations

Despite these challenges, some participants reported positive psychological changes. They expressed optimism about their future and personal goals, describing themselves as resilient and proactive. Hope was derived from the belief in recovery and the ability to prepare physically for surgery. One participant found joy in successfully adapting to new circumstances and developing inner strength. Gratitude was also a subtheme, with participants expressing appreciation for personal growth, family support, and the relatively limited physical impact of their condition, particularly the absence of life-threatening consequences. Participants described: *“Although we also had the support there, but I am, that’s what I mean, I am an optimistic person. I always try to be myself […].“And I think that also makes me strong. To believe in it too.” (ID11), or “And all in all, I would say that I started to feel more and more (.) a doctor who looked after me in the clinic also helped me a lot, so that I quickly developed a feeling of gratitude that it was just my hearing.” (ID1)*

#### 4.2.3 Theme 3: Social Changes

##### 4.2.3.1 Social interactions

Participants experienced strain on family and relationships due to their illness. Some felt they had become a burden, withdrew from social situations, or noticed friends distancing themselves, especially due to physical changes. Following the operation, ID7 explained that she had realized “*who of [her] friends was really there for [her] and who was not*”. She shared this experience with four other participants who reported a withdrawal from social contacts. In the case of ID4, the daughter had broken off contact with her, as ID4 had “*just sat there […] with a pained face*” when interacting with the grand-children, due to her hyperacusis. Since friends also withdrew, ID4 experienced profound social isolation:

*“And it took me ages to even realize it. What was happening to me. What was happening to my life. That you, that you become so (.) isolated.”*

ID7 also noted that her long-term partner reacted with shock to her physical changes. In everyday working life, ID2 also initially encountered confusion among his colleagues: “*You might take a short walk around the factory premises after (.) lunch and then I make a very strange movement, passing behind the person to get from the right side to the left side. It irritates most people at first and then you have to explain.”*

##### 4.2.3.2 Changes in social participation

Some participants struggled with social interactions, especially in noisy environments, and felt their illness was not always taken seriously. For example, ID1 said that the employment agency played down her physical impairments when she applied for severely disabled status: “*[…] which also brought me down a lot psychologically once again, because they simply then spun it that way that the whole problem wasn’t so blatant at the moment and I was just afraid that it would get so bad. And I always thought: “How fears that it’s going to get so bad?”. It’s kind of (.) I don’t hear anything now. What’s going to get worse? I didn’t feel seen at all […]”*

Some faced difficulties in employment, with a few reducing work hours or leaving their jobs. Work-place experiences varied, with some receiving support while others faced hostility. Many participants also had to cut back on leisure activities, including cultural events, sports, and group activities, due to pain and sensitivity to sound.

#### 4.2.4 Interaction of changes

Physical impairments often triggered psychological distress, including anxiety, frustration, and depressive symptoms. For example, hearing loss and tinnitus not only limited participants’ functional abilities but also increased emotional strain, particularly in social situations. Chronic pain and fatigue further exacerbated psychological challenges, reducing coping capacity and heightening stress sensitivity.

Psychological difficulties, in turn, affected social interactions. Participants reported withdrawal from family, friends, and work due to fear of stigma, frustration, or cognitive limitations. Emotional distress such as depression or anxiety often reinforced social isolation, creating a feedback loop that intensified overall burden.

Social changes also influenced both psychological and physical well-being. Negative reactions from colleagues, friends, or family contributed to feelings of alienation and reduced self-esteem, while supportive interactions facilitated adaptation and resilience.

Importantly, interactions with healthcare providers, such as perceived lack of understanding, insufficient information, or inadequate psychosocial support, further shaped patients’ psychological and social experiences. Positive and empathetic engagement from physicians, therapists, or support staff helped patients feel heard, validated, and supported, mitigating distress and improving overall adaptation.

Overall, the findings highlight a dynamic interplay between physical, psychological, and social domains, in which the role of healthcare providers can either buffer or amplify patients’ challenges. Challenges in one domain often triggered cascading effects in the others, underscoring the importance of a holistic, multidisciplinary approach to post-surgical care that addresses the interconnected nature of patients’ biopsychosocial experiences. Figure 1 provides a visual summary of these interactions. Figure 1 shows a summary of interactions of biopsychosocial changes as reported in this study.

**Figure 1.**
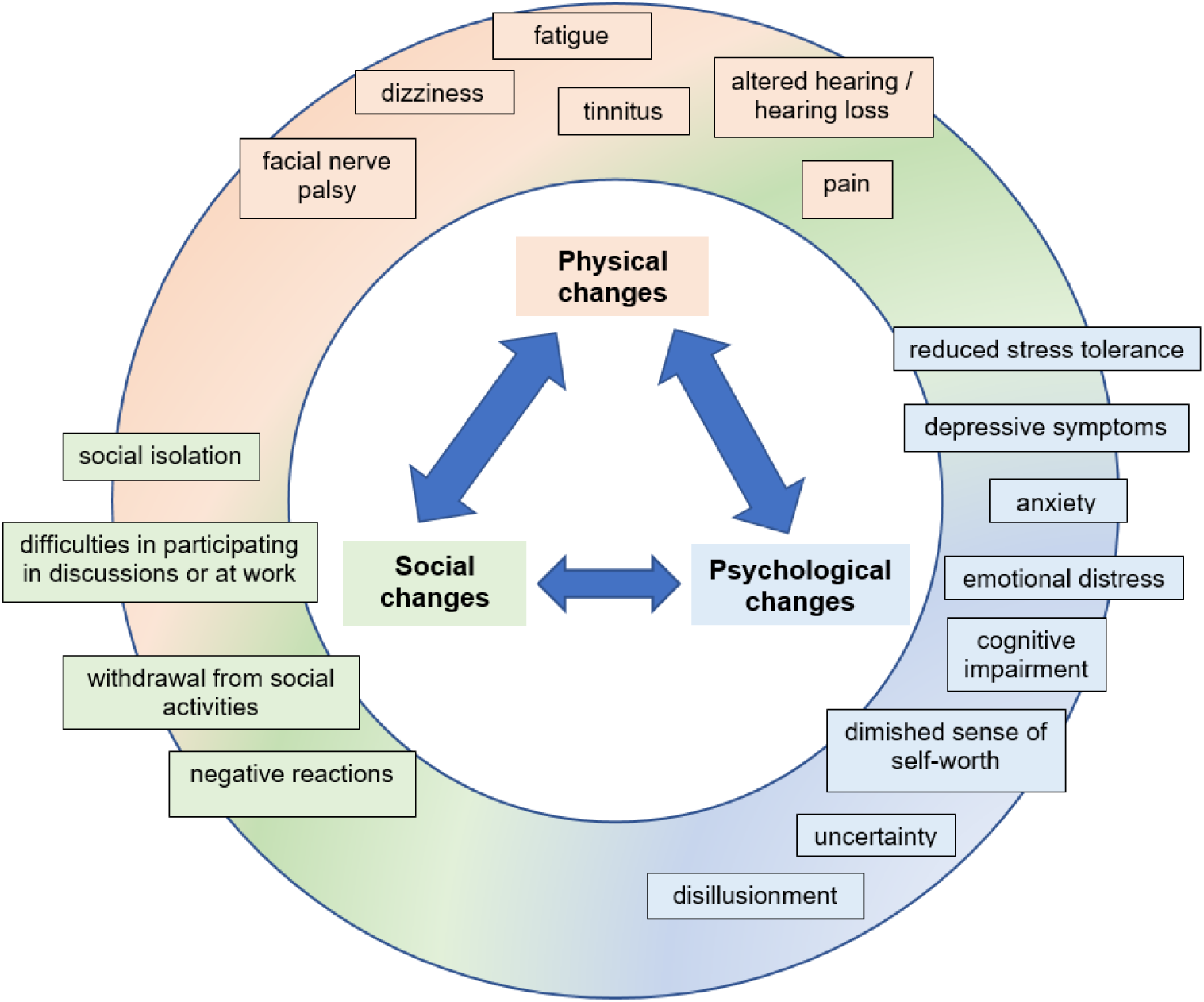
Interaction of biopsychosocial changes as reported by VS patients.

## 5. Discussion

### 5.1 Summary

Our study identified 16 subcategories across three main themes—physical, psychological, and social changes—demonstrating the multifaceted nature of the post-surgical experience for patients with VS. These categories interact, affecting various aspects of patients’ lives over the first five years following microsurgery.

Physically, altered hearing, dizziness, and pain were the most common issues. All participants experienced hearing impairment, with tinnitus and unilateral hearing loss causing significant emotional distress and social withdrawal as reported in other studies [14,15]. Dizziness and persistent balance problems were also frequent, consistent with findings on long-term vestibular dysfunction affecting quality of life [15,16]. Chronic pain, especially headaches contributed to frustration and despair, aligning with studies linking pain to psychological distress in VS patients [14,17,18].

Our findings highlight the interplay of physical symptoms with psychological and social factors. Participants reported how pain and stress worsened tinnitus and dizziness, creating a vicious cycle of distress. Feelings of being unheard by medical professionals increased frustration and helplessness, underscoring the need for better patient communication and psychological support [19]. Psychological interventions like cognitive-behavioral therapy and stress management could help mitigate these effects.

Psychologically, cognitive impairments, anxiety, and depressive symptoms were common. Concentration and memory difficulties affected many, consistent with reports that up to 20.3% of VS patients experience cognitive deficits post-surgery [15]. Anxiety was often related to social interactions and tumor recurrence [15,20]. Depressive symptoms, frequently linked to social isolation, were present in half of the sample, supporting findings that persistent vestibular symptoms increase depression risk [18]. Nevertheless, some participants also expressed optimism and gratitude, reflecting resilience seen in chronic illness [14,21].

Socially, hearing impairment impacted employment, with some reducing work hours or leaving jobs, paralleling findings that 14% of VS patients apply for disability benefits post-surgery [4]. Restrictions in leisure activities due to pain and sensory overload were common, consistent with research on sound sensitivity limiting social participation [14]. Communication difficulties, especially in noisy environments, posed major challenges, with VS patients struggling in group settings [15], a pattern also evident in our data. Importantly, our study highlights how social withdrawal can reinforce psychological distress, whereas strong social support appears to promote resilience. Participants who had access to peer-support groups or professional counseling described better adaptation, suggesting that structured psychosocial interventions could help mitigate isolation.

Our study builds on existing research by shedding light on the interconnected physical, psychological and social challenges that VS patients face after surgery. Physical symptoms, such as hearing loss, dizziness, pain, fatigue and facial nerve palsy, caused functional issues and triggered emotional distress. Hearing problems and tinnitus intensified anxiety in social situations, while chronic pain and fatigue contributed to depression and reduced stress tolerance. Visible impairments affected self-image, leading to insecurity and social anxiety. These psychological effects can influence social interactions, often causing patients to withdraw due to fear of stigma or cognitive difficulties. Emotional strain was further exacerbated by negative social reactions and dissatisfaction with medical care. In contrast, strong social support and open communication fostered resilience and adaptation. These findings are consistent with biopsychosocial models of chronic illness and emphasise the importance of holistic care (Figure 1).

Taking a holistic, multidisciplinary approach that combines medical, psychological and social support can greatly enhance long-term quality of life. It is important to inform VS patients early about the potential biopsychosocial effects to help them set realistic expectations. Many patients are unaware of the psychosocial changes that can occur after surgery and require psychological or social support. Practitioners can help by referring patients to self-help groups or specialists, such as psycho-oncologists and physiotherapists. Before patients return to their daily lives, it is important to assess whether they have adequate coping strategies and support in place, or if they require further care.

### 5.2 Strengths and Limitations

A major strength of this study was its in-depth qualitative design, which provided rich and nuanced insights into the long-term impact of VS microsurgery. Semi-structured interviews enabled participants to share their personal experiences, shedding light on the intricate interplay of physical, psychological and social factors. Focusing on long-term outcomes adds valuable perspective to a field that is often centred on short-term results.

However, several limitations must be noted. Recruiting participants via a self-help organisation may have introduced bias, as the participants tended to have higher education levels and be more engaged in health discussions. This may limit the applicability of the findings to individuals with lower health literacy. Additionally, the long time elapsed since surgery in many cases may affect recall accuracy and reduce the relevance of the study for patients in earlier phases of recovery. Finally, only including individuals who experienced significant post-surgical changes may have skewed the sample towards more negative outcomes. Future studies with broader sampling would help capture a fuller range of recovery experiences.

## 6. Conclusions

The findings of this study highlight the interconnectedness of physical, psychological, and social changes following VS microsurgery. Physical changes, such as hearing loss, dizziness, and pain, were frequently reported and contributed to psychological distress, including anxiety, depression, and cognitive impairments. For instance, hearing loss not only limited social interactions but also triggered feelings of frustration and isolation, which in turn exacerbated emotional well-being. These physical and psychological challenges then affected social participation, with many patients experiencing difficulties at work and in leisure activities. The interplay of these factors suggests the need for integrated care approaches that address both the physical and emotional aspects of recovery, helping patients better cope with the complex consequences of the surgery and improve overall health-related quality of life.

## Abbreviations

VS: Vestibular Schwannoma
hrQoL: health-related Quality of Life
VAN: Vereinigung Akustikus Neurinom e.V. (patient advocacy group)
IT: Interviewee (used to anonymize participants in quotes)
ID: Participant identifier (used in quotes, e.g., ID1, ID2)
Koos: Koos Classification (tumor size grading system)
COREQ: onsolidated Criteria for Reporting Qualitative Research

## Data Availability

All data produced in the present study are available upon reasonable request to the authors

## Acknowledgements

The authors would like to thank the members of the VAN, especially Mrs. Schaff and Mrs. Hühn for their support during the study. We would also like to thank all of the participants.

## Contribution list

MR is the responsible principle investigator of the study. NN was involved in planning and preparation of the study. NN recruited participants and collected data, and analysed the data with help of LB. All authors interpreted the results. MR wrote the first draft of the manuscript. SW, NN and IS critically revised the manuscript for important intellectual content. All authors gave final approval of the version to be published and agreed to be accountable for the work.

## Ethics statement

The study received approval from the local Ethics Institutional Review Board of the Psychosocial Center of the University Medical Center Hamburg-Eppendorf (LPEK-0694). All participants provided written informed consent prior to enrollment in the study.

## References

1. Carlson ML, Link MJ. Vestibular Schwannomas. N Engl J Med. 2021 Apr 8;384(14):1335–48.

2. Lucidi D, Fabbris C, Cerullo R, Di Gioia S, Calvaruso F, Monzani D, et al. Quality of life in vestibular schwannoma: a comparison of three surgical techniques. Eur Arch Oto-Rhino-Laryngol Off J Eur Fed Oto-Rhino-Laryngol Soc EUFOS Affil Ger Soc Oto-Rhino-Laryngol - Head Neck Surg. 2022 Apr;279(4):1795–803.

3. Bender M, Tatagiba M, Gharabaghi A. Quality of Life After Vestibular Schwannoma Surgery: A Question of Perspective. Front Oncol. 2021;11:770789.

4. Neve OM, Jansen JC, van der Mey AGL, Koot RW, de Ridder M, van Benthem PPG, et al. The impact of vestibular schwannoma and its management on employment. Eur Arch Oto-Rhino-Laryngol Off J Eur Fed Oto-Rhino-Laryngol Soc EUFOS Affil Ger Soc Oto-Rhino-Laryngol - Head Neck Surg. 2022 Jun;279(6):2819–26.

5. Hotchkies A, Heward E, Wadeson A, Heal C, Freeman SR, Rutherford SA, et al. Quality of Life Outcomes in Vestibular Schwannoma: A Prospective Analysis of Treatment Modalities. The Laryngoscope. 2025 Feb 27;

6. Alkins RD, Newsted D, Nguyen P, Campbell RJ, Beyea JA. Predictors of Postoperative Complications in Vestibular Schwannoma Surgery-A Population-Based Study. Otol Neurotol Off Publ Am Otol Soc Am Neurotol Soc Eur Acad Otol Neurotol. 2021 Aug 1;42(7):1067–73.

7. Thomas M, Führes H, Scheer M, Rampp S, Strauss C, Schönfeld R, et al. Perceived Health Benefits in Vestibular Schwannoma Patients with Long-Term Postoperative Headache: Insights from Personality Traits and Pain Coping-A Cross-Sectional Study. J Pers Med. 2024 Jan 8;14(1):75.

8. Thomas M, Rampp S, Scheer M, Strauss C, Prell J, Schönfeld R, et al. Premorbid Psychological Factors Associated with Long-Term Postoperative Headache after Microsurgery in Vestibular Schwannoma-A Retrospective Pilot Study. Brain Sci. 2023 Aug 7;13(8):1171.

9. Thomas M, Scheer M, Rampp S, Strauss C, Schönfeld R, Leplow B. Psychological factors and long-term tinnitus handicap in vestibular schwannoma patients after retrosigmoid microsurgery - a cross-sectional study. Int J Audiol. 2025 Mar;64(3):209–16.

10. Rutenkröger M, Scheer M, Rampp S, Strauss C, Schoenfeld R, Leplow B. Associations between Psychological Factors and Long-Term Dizziness Handicap in Vestibular Schwannoma Patients: A Cross-Sectional Study. 2024.

11. Pruijn IMJ, Kievit W, Hentschel MA, Mulder JJS, Kunst HPM. What determines quality of life in patients with vestibular schwannoma? Clin Otolaryngol Off J ENT-UK Off J Neth Soc Oto-Rhino-Laryngol Cervico-Facial Surg. 2021 Mar;46(2):412–20.

12. Tong A, Sainsbury P, Craig J. Consolidated criteria for reporting qualitative research (COREQ): a 32-item checklist for interviews and focus groups. Int J Qual Health Care J Int Soc Qual Health Care. 2007 Dec;19(6):349–57.

13. Kuckartz U. Qualitative Inhaltsanalyse. Methoden, Praxis, Computerunterstützung. 4. Auflage. Weinheim; Basel: Beltz Juventa 2018. (Grundlagentexte Methoden).

14. Brooker J, Burney S, Fletcher J, Dally M. A qualitative exploration of quality of life among individuals diagnosed with an acoustic neuroma. Br J Health Psychol. 2009 Sep;14(Pt 3):563–78.

15. Pruijn IMJ, van Heemskerken P, Kunst HPM, Tummers M, Kievit W. Patient-preferred outcomes in patients with vestibular schwannoma: a qualitative content analysis of symptoms, side effects and their impact on health-related quality of life. Qual Life Res Int J Qual Life Asp Treat Care Rehabil. 2023 Oct;32(10):2887–97.

16. Myrseth E, Møller P, Wentzel-Larsen T, Goplen F, Lund-Johansen M. Untreated Vestibular Schwannoma: Vertigo is a Powerful Predictor for Health-Related Quality of Life. Neurosurgery. 2006 Jul 1;59(1):67–76.

17. Osterhaus JT, Townsend RJ, Gandek B, Ware JE. Measuring the functional status and well-being of patients with migraine headache. Headache. 1994 Jun;34(6):337–43.

18. Rutenkröger M, Wandke S, Gempt J, Dührsen L, Scheer M, Strauss C, et al. German translation and cross-cultural adaptation of the Vestibular Schwannoma Quality of Life Index (VSQOL). J Patient-Rep Outcomes. 2024 Aug 13;8(1):95.

19. Ryzenman JM, Pensak ML, Tew JM. Patient perception of comorbid conditions after acoustic neuroma management: survey results from the acoustic neuroma association. The Laryngoscope. 2004 May;114(5):814–20.

20. Leong SC, Lesser TH. A United Kingdom survey of concerns, needs, and priorities reported by patients diagnosed with acoustic neuroma. Otol Neurotol Off Publ Am Otol Soc Am Neurotol Soc Eur Acad Otol Neurotol. 2015 Mar;36(3):486–90.

21. Ben-Harosh L, Barker-Collo S, Nowacka A, Garrett J, Miles A. Quality of life and broader experiences of those with acoustic neuroma: a mixed methods approach. Brain Impair [Internet]. 2024 Jan 25 [cited 2025 Feb 28];25(1). Available from: https://www.publish.csiro.au/ib/IB23072

